# Investigating the Role of Zinc in Mitigating Blood Lead Levels Toxicity on Gut Microbiome Diversity: NHANES 2007-2010

**DOI:** 10.1101/2024.10.31.24316539

**Authors:** Kathyrn R Ayres, Juan P. Liuzzi, Freeman C. Lewis, Huda M. Mobarki

## Abstract

**Introduction:** Zinc is an essential micronutrient used in many biological functions including maintaining the gut microbial diversity. Many environmental factors, such as lead exposure, have been shown to disrupt diversity. The purpose of this study was to determine whether zinc serves as a protective factor against elevated blood lead levels (BLL) on gut microbiome diversity.

**Methods:** The 2007-2008 and 2009-2010 NHANES datasets were utilized to conduct a cross-sectional complex survey analysis aimed at determining whether zinc intake acts as a protective factor against changes in microbiome diversity associated with BLL, using enterolactone (ENL) as a biomarker. A multiple linear regression was conducted to evaluate whether an interaction between BLL and zinc intake could predict ENL. The model included BLL, zinc intake and their interaction, along with additional covariates such as gender, fiber intake and BMI.

**Results:** BMI, fiber intake, and gender were identified as covariates through diagnostic analysis and stepwise regression and were included in the final model. Sequential variable selection revealed that fiber intake was a confounding variable in the relationship between zinc and ENL levels (p = 0.543), while gender was identified as a confounding factor between BLL and ENL levels (p = 0.173). After controlling for fiber intake, zinc intake was not significantly associated with predicting microbiome diversity (p = 0.101). Additionally, no significant interaction between zinc and BLL was observed in predicting ENL levels (p = 0.079).

**Conclusion:** Zinc intake did not play a crucial role in mitigating the toxicity of BLL exposure on gut microbiome diversity. However, the model did reveal important confounding variables, such as gender and fiber intake, which should be considered when using ENL as a biomarker. The public health implications suggest that dietary interventions focusing on fiber intake and managing BMI could be key in maintaining a diverse microbiome.

## 1. Introduction

### 1.1 Measuring Biodiversity

ENL is derived from plant lignans and have been proposed as a biomarker for measuring microbiome diversity.^1^ Ligands are metabolized by the gut microbiome into derivatives such as ENL and can be measured in the urine.^1^ Higher concentrations of ENL implies a more diverse gut microbial community. This is because their synthesis is only possible after the conversion of several other metabolites that require many species of bacteria in the gut microbiome to be present. ^1^

### 1.2 Microbiome

The gut is home to trillions of microorganisms that include bacteria, fungi, archaea, yeasts, protozoa, and viruses.^2^ Approximately 10^14^ organisms inhabit the microbiota and exist in a symbiotic relationship with the host.^2^ Each individual is colonized with a varying microbial ecosystem. A diverse population of microbiota is associated with a stable microbial environment and positive health outcomes for the host.^3^ Decreases in commensal bacteria diversity has been found in an array of medical conditions such as inflammatory bowel diseases,^3^ cognitive decline, emotional health issues,^4^ as well as metabolic and chronic diseases.^3^

This interaction with the host has a systemic impact on health status which is why research into the complex relationship between the gut microbiota and external stressors, such as environmental pollutants, is actively under investigation. Environmental pollutants, chemicals that cause disruption to biological pathways through the exposure of air, water, food, or soil, are one factor that has been identified as a cause of gut dysbiosis and inflammation.^5^ One group of environmental toxicants that have raised concerns of environmental and health experts are heavy metals.

### 1.3 Heavy metals

Heavy metals are chemically active in biological pathways.^6^ The absorption of toxic heavy metals, such as lead, can work to derail immunological functions and cause systemic oxidative stress in tissue. This can result in devastating impairment of physiological functions, including the gut. Toxic heavy metals that stay in the microenvironment of the gut disrupt the complex metal-microbe-host interplay.^6^ This disruption to homeostasis has been linked to alterations in the gut microbiome community and its diversity.^7^

Yet not all heavy metals are toxic. With a specific density of 7.14 g/cm^3^, zinc is considered a heavy metal.^8^ Unlike noxious heavy metals, zinc functions as an essential micronutrient in living organisms ranging from complex animal species to simplistic microbes.^9^ Twenty percent of the dietary intake of zinc is utilized by microbial bacteria.^10^ Zincs’ role in maintaining diversity is most likely linked to its ubiquitous nature. Bacteria utilize multiple zinc dependent mechanisms including modulating host defense, antioxidant systems, gene expression, and inhibiting transport of growth-promoting factors.^11^ Zinc has been shown to be essential in DNA repair, enzymatic reactions, responses to oxidative stress, four different transport systems, and regulatory roles in other physiological processes within the microbiome.^12^ Based on the harmful effects that lead has on the gut, including its disruption of bacterial communities, and zinc’s involvement in critical pathways within the microbiome, it is plausible that zinc may protect against lead’s detrimental effects on gut diversity.

### 1.4 Significance

Understanding the functionality of the gut microbiome is critical due to its systemic impact on health. Laboratory studies have confirmed that heavy metal exposure, including lead, reduces gut microbiome diversity.^13^ The present population study is the first to explore the potential protective role of zinc in maintaining this diversity at BLL found in the American population. Zinc may act as an effect modifier, mitigating lead’s negative impact on microbiome diversity due to its essential role in numerous biological pathways and its involvement in many taxa of gut bacteria. Additionally, using ENL as a biomarker for microbiome health in this context adds a novel layer to the research, providing a new avenue for understanding the interaction between zinc and lead in relation to gut health.

## Research Methods

### 4.1 Data

Data from the CDC’s NHANES 2007 - 2008 and 2009 - 2010 cycles were used to conduct a cross-sectional analysis. The NHANES datasets, which are based on a cross-sectional study design, employ a multistage, stratified, and clustered sampling approach with survey weights to ensure accurate representation of the American population. These methods were accounted for in the data analysis of this study. Zinc intake was assessed through 24-hour dietary recall interviews, where participants provided detailed reports of their food and beverage consumption. The USDA’s Food and Nutrient Database for Dietary Studies was used to calculate the nutrient content, including daily zinc intake, based on these records. Blood lead levels (BLL) were measured using inductively coupled plasma mass spectrometry (ICP-MS), a highly accurate method for detecting lead concentrations in whole blood. Microbiome diversity was indirectly assessed through the measurement of ENL, a metabolite produced by gut bacteria, which was collected via urine samples. These data collection methods allowed for a robust analysis of the relationship between zinc intake, BLL, and gut microbiome diversity.

Zinc intake, fiber intake, BLL, BMI ENL, age, gender, and creatinine were merged and uploaded into SPSS. ENL was collected in cycles between 1999 – 2010 and supplement intake was collected starting in the 2007-08 cycle. Including supplement intake was deemed important to capture accurate zinc and fiber intakes. This limited the study to the 2 cycles. The two 24-hour recalls for zinc food and supplement intake were both added and averaged to determine total zinc intake. Missing data from food intake was replaced by the means for each cycle. Carry over imputation was used to avoid non-response bias for missing data for supplements because not every participant used supplements. The same was done for fiber intake. Individuals below the age of 18 and older than 50 were removed due to the changes in microbiome diversity at these stages of life.^14^ ENL was adjusted for creatinine levels by dividing ENL, measured in ng/dL, by creatinine, measured in mg/dL, resulting in units of ng/mg/dL. This adjustment corrects for differences in urine dilution among individuals.^1, 15^ ENL was also common log transformed to account for its non-normal distribution. Lead levels were measured in the blood in µg/dL.

### 2.2 Complex Survey Analysis

A complex survey design was accounted for using RStudio’s Survey Package in the analysis. Descriptive statistics were run for all variables. Following diagnostic testing, a forward stepwise regression approach was employed to assess the relationships between the covariates and independent variables, focusing on their predictive capacity for ENL levels. An interaction term was incorporated to evaluate whether varying levels of zinc and BLL can predict ENL levels. The final model included BLL and zinc as independent variables, and accounted for fiber intake, BMI, and gender as covariates as well the zinc and BLL interaction term. All statistical analyses and visualizations were performed using RStudio (Version 2023.09.1+494) accessed through Anaconda.

## 3. Results

### 3.1 Descriptive

After removing missing values, the study included a total of 1,887 participants, with an even distribution of females (51%) and males (49%) (Table 1). Continuous variables revealed all participants had BLL in their system, with BLL concentrations ranging from a minimum of 0.18 µg/dL to a maximum of 11.8 µg/dL and an average of 1.24 µg/dL. Participants’ ages ranged from 18 to 48 years, with a mean age of 33.47 years (Table 2). The analysis revealed significant gender differences in several variables. Females had significantly higher levels of ENL than males, with a mean difference of 0.25 (p-value = <0.001). This suggests that females may exhibit greater gut microbiome diversity compared to males. The analysis also found a substantial gender difference in BLL, where males had significantly higher BLL than females, with a mean difference of 0.59 µg/dL (p-value = <0.001). Although there was a significant difference in zinc, this is logical as women’s RDA is 8 mg/day and men’s is 11 mg/day. Similarly, males consumed significantly more fiber than females, with a mean difference of 2.82 grams (p-value = 2.00e-06). Unlike zinc, fiber intake was below the RDA for both men and women (38 grams/day for men and 25 grams/day for women), with average intakes of 17.81 grams/day for men and 14.99 grams/day for women. Both men and women’s average BMI was considered overweight and did not show a significant difference between genders (Table 3).

**Table 1:** Study Participants.

| Gender | Proportion | n |
| --- | --- | --- |
| Male | 0.49 | 925 |
| Female | 0.51 | 962 |
| Total | 1.0 | 1887 |

**Table 2:** Descrritive statistics.

| Variable | Weighted Mean | Weighted SD | Max | Min |
| --- | --- | --- | --- | --- |
| Log10_ENL | 2.28 | 0.81 | 4.47 | -0.93 |
| BMI (kg/m<sup>2</sup>) | 28.16 | 7.03 | 68.63 | 15.49 |
| AGE (years) | 33.47 | 8.99 | 48 | 18 |
| Total Fiber (g/day) | 16.36 | 8.09 | 68.75 | 1.1 |
| Total Zinc (mg/day) | 16.08 | 9.79 | 113.02 | 1.05 |
| BLL (μg/dL) | 1.24 | 0.99 | 11.8 | 0.18 |

**Table 3:** Gender specific descriptive statistics.

| Variable | t-statistic | df | p-value | Male: Mean (SE) | Female: Mean (SE) |
| --- | --- | --- | --- | --- | --- |
| Log10_ENL | 5.1724 | 31 | < 0.0001 | 2.147084 (0.0354) | 2.400567 (0.030) |
| Total Fiber (g/day) | -5.829 | 31 | < 0.0001 | 17.80844 (0.488) | 14.98591 (0.307) |
| Total Zinc (g/day) | -4.0465 | 31 | 0.0003 | 17.23821 (0.534) | 14.98044 (0.410) |
| BMI (kg/m<sup>2</sup>) | -0.2622 | 31 | 0.7949 | 28.26989 (0.295) | 28.17684 (0.278) |
| BLL | -11.838 | 31 | < 0.0001 | 1.55433 (0.053) | 0.965175 (0.022) |

**Table 4:** Forward Stepwise Regression Model Selection with AIC.

| Step | Variable | Coefficient | SE | t-value | p-value | AIC |
| --- | --- | --- | --- | --- | --- | --- |
| 1 | Intercept | 2.200588 | 0.039038 | 56.37 | < 0.001 | 4340.13 |
|  | Total Zinc | 0.00527 | 0.002232 | 2.361 | 0.025 |  |
| 2 | Intercept | 2.266742 | 0.041158 | 55.075 | < 0.001 | 4334.26 |
|  | Total Zinc | 0.005365 | 0.002232 | 2.403 | 0.023 |  |
|  | Lead | -0.05464 | 0.020815 | -2.625 | 0.014 |  |
| 3 | Intercept | 2.853902 | 0.098997 | 28.828 | < 0.001 | 4279.94 |
|  | Total Zinc | 0.004558 | 0.002305 | 1.978 | 0.056 |  |
|  | Lead | -0.06095 | 0.019025 | -3.203 | 0.003 |  |
|  | BMI | -0.02012 | 0.002849 | -7.061 | < 0.001 |  |
| 4 | Intercept | 2.660036 | 0.0938 | 28.359 | < 0.001 | 4244.87 |
|  | Total Zinc | 0.00595 | 0.002354 | 2.527 | 0.017 |  |
|  | Lead | -0.0275 | 0.019667 | -1.398 | 0.173 |  |
|  | BMI | -0.01989 | 0.002789 | -7.132 | < 0.001 |  |
|  | GENDER2 | 0.241185 | 0.050048 | 4.819 | < 0.001 |  |
| 5 | Intercept | 2.740478 | 0.098501 | 27.822 | < 0.001 | 4270.35 |
|  | Total Zinc | 0.001632 | 0.002653 | 0.615 | 0.543 |  |
|  | Lead | -0.06335 | 0.018641 | -3.399 | 0.002 |  |
|  | BMI | -0.01957 | 0.002779 | -7.042 | < 0.001 |  |
|  | Total Fiber | 0.009044 | 0.003342 | 2.706 | 0.011 |  |
| 6 | Intercept | 2.42935 | 0.104723 | 23.198 | < 0.001 | 4226.4 |
|  | BMI | -0.019 | 0.002656 | -7.152 | <0.001 |  |
|  | GENDER2 | 0.266067 | 0.052903 | 5.029 | <0.001 |  |
|  | Total Fiber | 0.01139 | 0.003406 | 3.344 | 0.003 |  |
|  | Lead | 0.023971 | 0.03494 | 0.686 | 0.499 |  |
|  | Total Zinc | 0.006034 | 0.00355 | 1.7 | 0.101 |  |
|  | Zinc: Lead | -0.00293 | 0.001604 | -1.826 | 0.079 |  |

### 3.2 Multiple Linear Regression

Table 3 shows a total of 6 regression models were created in the forward stepwise process. The initial model only considered zinc intake’s ability to predict ENL. Zinc showed a statistically significant positive relationship with ENL (p = 0.025). This indicates that higher levels of zinc are associated with increased ENL levels, albeit the effect is modest. The second model included lead and zinc. Model 2 identified a significant negative relationship between BLL (p = 0.0135). This suggests that higher BLL are associated with lower ENL levels, supporting the hypothesis that lead exposure may adversely affect microbiome diversity. In Model 3, BMI emerged as a significant predictor (p < 0.001), indicating that higher BMI is associated with lower ENL levels. The inclusion of gender in Model 4 revealed a significant positive relationship (p = <0.001). However, the addition of gender resulted in the loss of significance in BLL. In model 5 fiber replaced the addition of gender as a covariate. Upon its addition zinc lost its significance. Finally in model 6, the interaction term between BLL and zinc was included in the model with all of the covariates. The interaction term between BLL and zinc exhibited a p-value of 0.079, indicating that while it did not reach conventional significance, it is close to the threshold.

Akaike Information Criterion (AIC) was used to determine how well the model explains variation in ENL because of the studies complex survey design. Unlike R^2^, which can misleadingly increase with additional variables and does not adequately account for the design effects associated with sampling methods and stratification, AIC helps identify the most effective predictors by penalizing overfitting. In this study, the AIC values ranged from 4226.40 to 4340.13. The decreasing AIC values across models suggest that adding variables improves the model’s fit. With the exception of the addition a fiber. However, since AIC is used to compare models rather than measure explained variance, it does not indicate how much of the variation in ENL levels is accounted for. This suggests that other unmeasured factors may still influence ENL levels, warranting further investigation.

## 4. Discussion

### 4.1 Enterolactone as a biomarker

ENL has been used as a biomarker in research but not as a standard protocol in clinical settings. However, both human and population studies have found reliability with this biomarker. A human study published by (Hullar et al, 2015) investigated the association between the diversity of the microbiome and presence of ENL in premenopausal women.^1^ Urine samples were used to quantify ENL and stool samples were obtained to measure diversity in the gut microbiome using 16S rRNA.^1^ They concluded metabolism of lignans in the gut is dependent on the presence of a diverse community of microorganisms to perform the series of reactions.^1^ A review published by (Lampe, 2003) even predicted that advancements in methodologies for measuring lignans will result in the use of their derivatives as biomarkers in large population-based studies.^16^

The analysis from this current population study supports that ENL is a reliable biomarker for gut microbiome diversity. The regression model found a positive correlation between fiber and ENL. Fiber intake has shown to increase diversity in the gut microbiome.^17^ Inversely, increased BMI, related to obesity, has been associated with decreased diversity in the gut microbiome.^18^ BMI was found to have a significantly negative correlation with ENL in the regression model.

### 4.3 Confounding factors

When total fiber was added to the model, the significance of zinc was lost, with the coefficient estimate for zinc changing from 0.004558 to 0.001632. This represents a percentage change of approximately 64.25%, indicating that zinc may not have a meaningful impact on ENL levels when fiber is accounted for. The substantial change in the estimate suggests that zinc’s apparent influence on ENL was likely an overestimation prior to including fiber in the model, highlighting the importance of considering confounding factors like dietary fiber in future analyses.

When gender was incorporated into model the estimate for lead changed from −0.06095 to −0.02750, a 54.9% increase, and its significance diminished from (p = 003) to (p = 0.1730). The adjustment for gender revealed a need to account for it as a confounding factor in understanding its influence on ENL levels. Diagnostic tests found that men have significantly higher BLL and lower ENL compared to women. While neither men or women met their fiber intake needs on average, it is possible men’s higher exposure to lead coupled with low fiber intake could also contribute to reduced ENL levels. This highlights a complex relationship between the covariate, independent and dependent variables.

### 4.2 Zincs’ role in protecting diversity

In a linear regression, an interaction between zinc intake and BLL would have suggested that the effect of lead on the outcome (e.g., ENL levels) depended on the level of intake zinc. Specifically, if zinc mitigated the harmful effects of lead, the interaction term would reflect that zinc alters the relationship between lead and the outcome. Although the data did not support a significant interaction between zinc and BLL in their effect on ENL, further investigation may still be warranted. The interaction term was estimated at 0.079, close to the threshold of significance. Future studies could explore other models, such as an ANCOVA, by categorizing zinc and BLL. Additionally, a sensitivity analysis may help identify the biological thresholds at which changes in ENL occur, particularly in response to specific levels of zinc intake or lead toxicity.

### 4.4 Weakness

There were several weaknesses of this study to consider. Firstly, this study used self-reported 24-h recalls which may not be as accurate as direct reporting. Also, diversity is influenced by many variables that were not included in this model such as genetics, disease, mode of delivery, infant feeding, medication, exposure to other pollutants and dietary factors.^19^ The current study included only three covariates and had a high AIC. This supports that other unmeasured factors may significantly influence the outcomes, warranting further exploration.

Literature supports that low intake of zinc causes a decrease in diversity through deficiency and high intake causes a decrease in diversity related to toxicity.^20^ A review published by (Cheng et al., 2023) found a decrease in α-diversity in rat, mice, pigs, and broiler chicken models following zinc deficiency and overload.^21^ The majority of the participants in the present study had optimal zinc intake, meaning these extreme zinc intakes were rare in the data. Also, lead toxicity is low America. This prevented the true relationship of a dose response curve to be fully analyzed and helps to explain the opposing results from other studies. One way to address this would be to utilize data from a population that has higher lead exposure and zinc deficiency rates.

Diversity was measured by the concentration of the ENL metabolite which is synthesized after a series of reactions requiring many species of bacteria. This gives a snapshot into the changes in diversity. However, measuring changes in diversity is complex. For a more comprehensive understanding of changes in diversity, bacteria populations are identified through 16s rRNA sequencing and analyzed through alpha and beta diversity analysis. Alpha diversity considers richness, evenness, and phylogenic diversity and beta diversity uses distant metrics to measure communal changes at different taxological levels. ENL is limited to representing only a subset of the microbial community dynamics and may not be accurately capturing the changes in bacterial populations. This is important because the removal of heavy metals from the gut seems to rely on the presence of specific bacteria species.^22, 23^

## 5. Conclusion

It was hypothesized that effect modification would be caused by zinc in the relationship between BLL and ENL, however this was not supported in the current analysis, as the interaction between zinc and BLL was not statistically significant. In fact, due to confounding relationships neither zinc nor BLL were found to significantly predict ENL levels in the model. Zinc’s potential impact on bacterial diversity may not have been fully captured in this design, and future studies should explore different models, such as ANCOVA and sensitivity analysis, from data with a case control design to better understand these interactions. The data did support that including factors like gender and fiber intake when using ENL as a biomarker is important, as they may confound the relationship between independent and dependent variables. Given discovery that lead may be more toxic than once thought, and the gut microbiome’s influence on immune function, nutrient absorption, and inflammation regulation, further investigation is recommended.^28, 29^

## Data Availability

The data utilized in this analysis was sourced from the open dataset provided by the National Health and Nutrition Examination Survey (NHANES), available through the Centers for Disease Control and Prevention (CDC) website. The cleaned data was uploaded as supplementary materials.

https://www.cdc.gov/nchs/nhanes/index.htm

